# Extension of the FUNC score for prediction of 12-month functional independence after primary intracerebral hemorrhage

**DOI:** 10.64898/2026.05.27.26354249

**Authors:** Joel Neves Briard, Vedant Kansara, Qi Shen, You Lim Song, Amy B. Cami, Angela G. Velazquez, Justin M. Esposito, Alex J. Klein, Shivani Ghoshal, Sachin Agarwal, Soojin Park, E. Sander Connolly, David Roh, Jan Claassen

**Author notes:** **Correspondence to:** Dr Jan Claassen, Columbia University Irving Medical Center, 177 Fort Washington Avenue, Milstein Building 8GS-300, New York, NY 10032, USA, Facsimile.

## Abstract

**Background:** The Functional Outcome in Patients with Primary Intracerebral Hemorrhage (FUNC) score was initially validated for prediction of functional independence on the Glasgow Outcome Scale (GOS) 90 days after intracerebral hemorrhage (ICH), but recovery often extends beyond three months.

**Aims:** Our objective was to extend the FUNC score for prediction of 12-month functional independence to strengthen its utility for family counseling and research methodology.

**Methods:** We conducted a single-center prospective cohort study enrolling adult patients with primary ICH between February 2009 and January 2018. We calculated FUNC scores at admission and assessed GOS 12 months after ICH. The primary outcome was 12-month functional independence, defined as a GOS score ≥4. We calculated the area under the receiver operating characteristic curve (AUC) of the FUNC score using logistic regression, handling missing GOS with multiple imputation by chained equations. We evaluated score calibration using a calibration curve and the Brier score, and we assessed clinical utility using decision curve analysis. We explored the statistical efficiency gains of using FUNC-based sliding dichotomy thresholds for favorable outcome definitions by running simulations of a clinical trial with 1:1 randomization. We ran 5000 simulations for each sample size (100 to 1000, in increments of 10) and treatment effect (odds ratio of 1.5, 2.0 and 2.5) combination and calculated efficiency gains for each respective treatment effect as the percentage reduction in sample size required to have 80% power using sliding versus fixed dichotomy thresholds.

**Results:** A total of 535 patients were included (median [IQR] age 68 [54-79], 237 [44%] female, median [IQR] NIHSS 16 [6-25], median [IQR] FUNC 8 [6-9]). Overall, 99 of 445 (22%) patients with known 12-month GOS achieved functional independence. The FUNC score had an AUC of 0.79 (95%-CI: 0.75-0.84) for 12-month functional independence. The calibration plot was reasonable, with modest evidence of overestimation at low predicted probabilities, and the Brier score was 0.15. A net benefit was observed across 5-50% threshold probabilities. Sliding dichotomy had an efficiency gain of 27% for a treatment effect of OR=2.0, and a gain of 22% for a treatment effect of OR=2.5. The efficiency gain for a treatment effect of OR=1.5 could not be calculated because the fixed dichotomy did not reach 80% power despite a sample size of 1000 patients.

**Conclusions:** The FUNC score’s predictive performance for 12-month functional independence was comparable to its originally validated 3-month discrimination. Following external validation across centers, the FUNC score may be leveraged to counsel families on global measures of long-term functional independence and to implement sliding dichotomy methodology in ICH research.

## INTRODUCTION

Despite advances in acute medical and surgical management, primary intracerebral hemorrhage (ICH) remains a highly morbid and frequently fatal condition.^1^ Several prognostic scales to predict long-term functional recovery have emerged in recent years, but many lack sufficient internal validity or robust external validation — limiting their endorsement by expert societies.^2–4^ This gap is especially consequential in ICH patients who develop disorders of consciousness such as coma, where perceptions of poor prognosis can drive early withdrawal of life-sustaining therapies (WLST).

The FUNC score (Functional Outcome in Patients with Primary Intracerebral Hemorrhage) was developed to estimate the probability of functional independence at 90 days using clinical data routinely available at hospital admission.^5^ However, recovery after ICH extends beyond three months, underscoring the need for prediction tools with longer time horizons.^6, 7^ Several recent large-scale ICH trials defined 6- or 12-month functional outcomes as their primary endpoint, reflecting growing consensus that three months is too early to capture the full treatment effect in this patient population.^8–10^ Extending the FUNC score to 12-month functional independence after ICH could therefore strengthen its utility for both family counseling and clinical research. The primary objective of this study was to evaluate the prognostic accuracy of the FUNC score for 12-month functional independence on the GOS in patients with acute primary ICH. The secondary objective was to explore the statistical efficiency gains of FUNC-based sliding dichotomy methodology using a clinical trial simulation.

## METHODS

This is a secondary analysis of the Intracerebral Hemorrhage Outcomes Project (ICHOP), a prospective observational cohort study of individuals with acute ICH admitted to the Columbia University Irving Medical Center.^11^ We included patients enrolled in ICHOP between February 2009 and January 2018. ICHOP was approved by our Institutional Review Board (IRB-AAAD4775) and informed consent was obtained from each participant’s legal representative. Study reporting follows guidance from the TRIPOD+AI statement.^12^ The data and code that support the findings of this study are available from the corresponding author upon reasonable request.

### Study population

The study population comprises adults with acute primary ICH. We consecutively enrolled individuals 18 years or older with an admission diagnosis of primary ICH supported by evidence of an intraparenchymal hematoma on brain CT or MRI scan. We excluded patients with primary intraventricular hemorrhage, ICH secondary to trauma or vascular lesions such as arterio-venous malformations, hemorrhagic conversion of an ischemic stroke or cerebral malignancy, and multiple acute brain injury causes. Trained research staff monitored daily ICH admissions and approached patients and their surrogate decision makers, when pertinent, for enrollment.

### Study procedures

We prospectively collected baseline characteristics at enrollment. Trained research coordinators extracted clinical data from medical charts, including age, sex, premorbid modified Rankin score (mRS), history of cognitive impairment, admission Glasgow Coma Scale (GCS), and admission National Institutes of Health Stroke Scale (NIHSS). They extracted neuroimaging data from the initial brain scans, including localization of the hematoma and presence of intraventricular hemorrhage, and they calculated hematoma volumes using Medical Image Processing, Analysis, and Visualization software (National Institutes of Health; https://mipav.cit.nih.gov/index.php). The admission FUNC score was calculated based on age, premorbid cognitive impairment, hematoma location, hematoma volume, and admission GCS (**Table S1**). FUNC scores could range from 0 to 11, with higher scores indicating higher probabilities of functional independence at 90 days.^5^

### Outcomes

Trained assessors obtained GOS scores by a structured telephone interview three, six, and 12 months after ICH. Post-hospitalization outcomes were obtained from patients whenever possible, and from close relatives or caregivers if patients were unable to communicate. The primary outcome was functional independence, defined as a GOS score ≥4, 12 months after ICH. Secondary outcomes were functional independence at three and six months. We recorded additional hospitalization outcomes, including in-hospital death, WLST, hospital discharge mRS and disposition.

### Statistical analysis

Descriptive statistics are provided for the entire cohort, with continuous variables reported as medians and interquartile ranges and dichotomous variables as counts and proportions. To assess for potential bias introduced by differential loss to follow-up, patients with and without 12-month GOS data were compared using standardized mean differences. To account for missing 12-month GOS data, we performed multiple imputation by chained equations. The imputation model included FUNC score variables and other variables identified *a priori* as potentially associated with loss to follow up (sex, premorbid mRS, discharge mRS, discharge disposition, WLST, and 3- and 6-month GOS). We created 15 imputed datasets based on the observed proportion of missing 12-month GOS values.^13, 14^

In primary analyses, the discriminative performance of the FUNC score for predicting 12-month functional independence was assessed using logistic regression with the FUNC score as the sole predictor. Model discrimination was quantified using the area under the receiver operating characteristic curve (AUC). The model was evaluated using 10-fold cross-validation within each imputed dataset, and within-imputation AUC variances were derived via bootstrap resampling. Results were pooled across imputed datasets using Rubin’s Rules, with total variance decomposed into within- and between-imputation components.^15^ Calibration was assessed by plotting a calibration curve and calculating a pooled Brier score.^16^ Clinical utility was assessed using decision curve analysis, computing net benefit at threshold probabilities between 5% and 50% within each imputed dataset and averaging across imputations.^17^

In sensitivity analyses, we performed a complete case analysis by applying the logistic regression model to patients with known 12-month GOS. The model was again evaluated using 10-fold cross-validation to calculate an AUC, for which a 95% confidence interval was derived by bootstrap resampling. In exploratory subgroup analyses, we assessed the FUNC score’s predictive discrimination according to age (≤70 versus >70), sex (male versus female), ICH location (lobar versus deep versus infratentorial), ICH etiology (hypertensive microangiopathy versus cerebral amyloid angiopathy versus all other etiologies), and admission GCS (≤8 versus >8).

In exploratory secondary analyses, we tested the suitability of the FUNC score as a prognostic stratification instrument for a sliding dichotomy outcome framework in future ICH trials. We first derived prognostic strata from the complete-case cohort using tertiles of the predicted probability of 12-month functional independence generated by a logistic regression model of the FUNC score. Within each stratum, we identified the GOS threshold that maximised equipoise, defined as the observed favorable outcome probability closest to 50%. The statistical efficiency of the sliding dichotomy relative to fixed dichotomy was evaluated through a simulation study. For each combination of total sample size (N = 100 to 1000, increments of 10) and assumed treatment effect (odds ratios [OR] of 1.5, 2.0, and 2.5), 5000 trial datasets were generated under 1:1 randomisation. Patients were allocated to prognostic strata according to the empirical stratum proportions observed in the cohort. Control arm outcome probabilities were set to the observed probabilities of a favorable outcome within each stratum at the assigned GOS threshold. Treatment arm probabilities were derived by applying the assumed OR as a uniform log-odds shift from the control arm probability, consistent with a proportional odds assumption across strata. Binary outcomes (favorable or unfavorable outcome) were simulated using a binomial distribution. Treatment and control arm outcomes were compared across prognostic strata using the Cochran-Mantel-Haenszel test, with a two-sided α of 0.05. Power was estimated as the proportion of simulated trials achieving statistical significance across 5000 iterations. The sample size required to achieve 80% power was identified as the smallest number of patients at which the power curve crossed the 0.80 threshold for each outcome framework and assumed treatment effect. We calculated efficiency gains for each respective treatment effect as the percentage reduction in sample size required to have 80% power using sliding versus fixed dichotomy thresholds. Analyses were performed in R, version 4.5.1 (Vienna, Austria).

## RESULTS

Among 642 patients enrolled in ICHOP, 535 were included in the study (**Figure 1**). Baseline characteristics are detailed in **Table 1**. Median (interquartile range [IQR]) age was 68 (54-79) years; 237 (44%) participants were female. At admission, median (IQR) NIHSS was 16 (6-25) and median (IQR) FUNC was 8 (6-9). At follow up, 73 (14%), 99 (19%) and 90 (17%) patients had missing 3-, 6-, and 12-month GOS, respectively. Characteristics of 445 patients with known 12-month GOS and 90 patients lost to 12-month follow up are compared in **Table S2**: high standardized mean differences in some baseline and hospital outcome characteristics signaled data missingness was not completely at random. Distributions of GOS scores at 3, 6 and 12 months after ICH are displayed in **Figure 1**. At 12 months, 99 of 445 patients with known GOS (22%) had achieved functional independence. The proportions of patients who achieved 12-month functional independence ranged from 3% in patients with a FUNC score of 0-4 to 74% in patients with a FUNC score of 11 (**Figure S1**).

**Figure 1.**
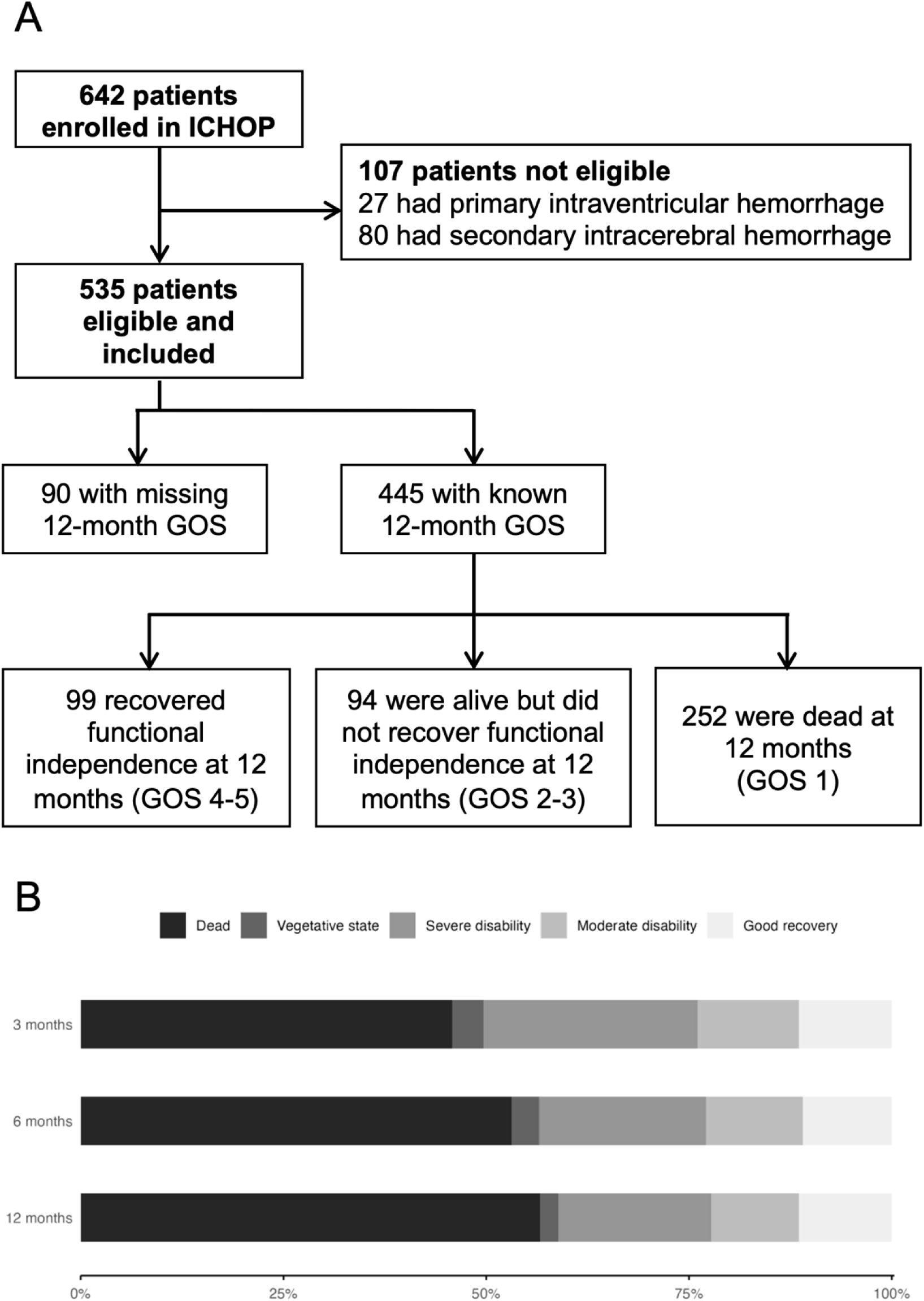
Flowchart diagram and distributions of GOS scores at 3, 6 and 12 months. **Panel A:** Flowchart diagram. **Panel B:** Distributions of 3-, 6- and 12-month GOS scores. ICHOP denotes Intracerebral Hemorrhage Outcomes Project and GOS denotes Glasgow Outcome Scale.

**Table 1.**
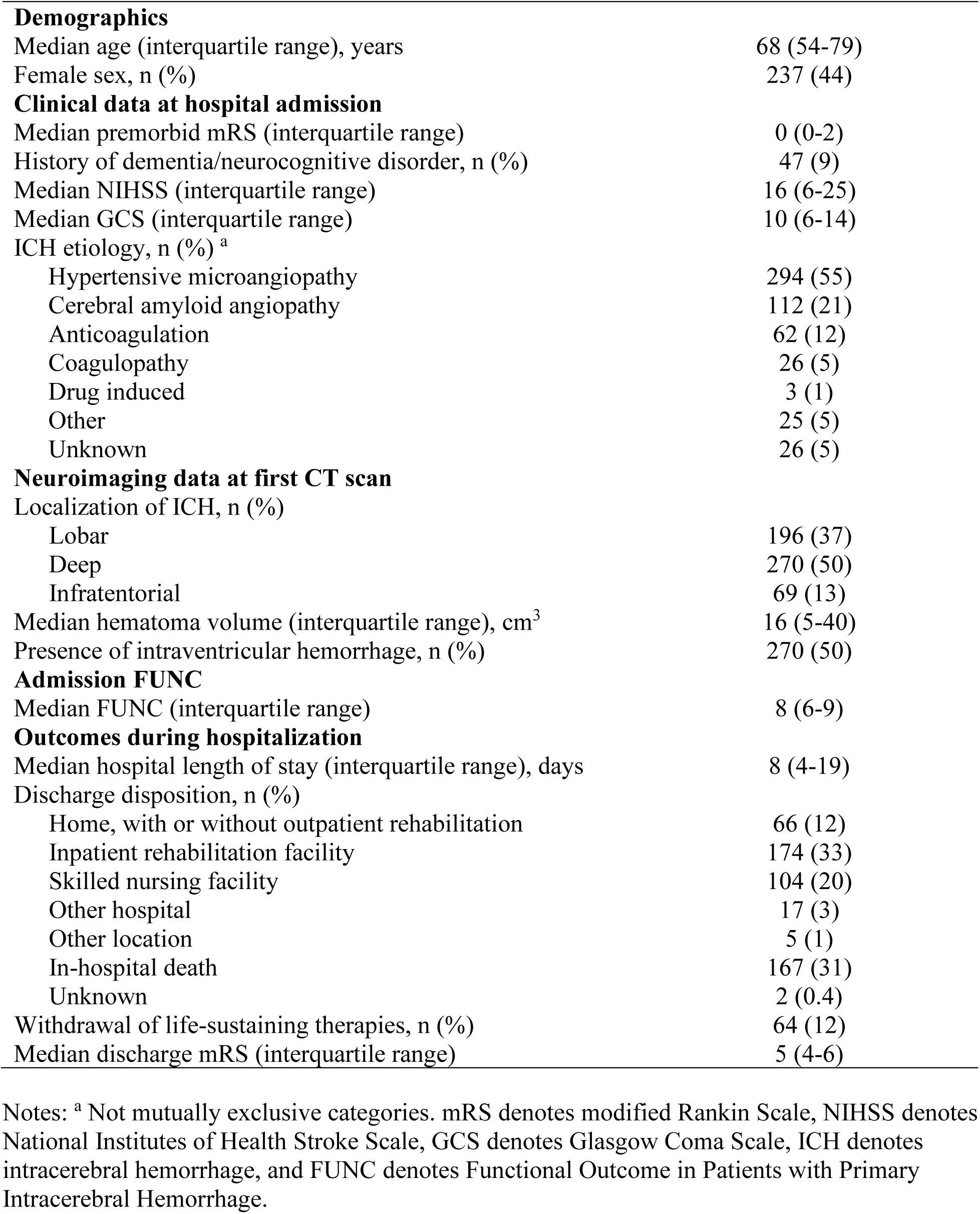
Baseline participant characteristics and in-hospital outcomes.

### Discrimination, calibration and decision curve analysis

Results from primary analyses are provided in **Figure 2**. The FUNC score had an AUC of 0.79 (95%-CI: 0.75-0.84) for 12-month functional independence. The score showed reasonable calibration in the mid-range of predicted probabilities, with discrete evidence of overestimation at low predicted probabilities (∼<20%) and underestimation at moderate predicted probabilities (∼20-50%; Brier score: 0.15). Decision curve analysis demonstrated that the FUNC score provided greater net benefit than treat-all and treat-none strategies across 5-50% threshold probabilities. Results were comparable in the complete case sensitivity analysis, where the FUNC score had an AUC of 0.81 (95%-CI: 0.77-0.86) and showed similar calibration (Brier score: 0.13, **Figure S2**). In secondary analyses, the FUNC score showed comparable discrimination and calibration for 3-month (AUC: 0.78, 95%-CI: 0.74-0.83; Brier score: 0.15) and 6-month functional independence (AUC: 0.79, 95%-CI: 0.74-0.84; Brier score: 0.16). FUNC score discrimination across subgroups is provided in **Figure 3**.

**Figure 2.**
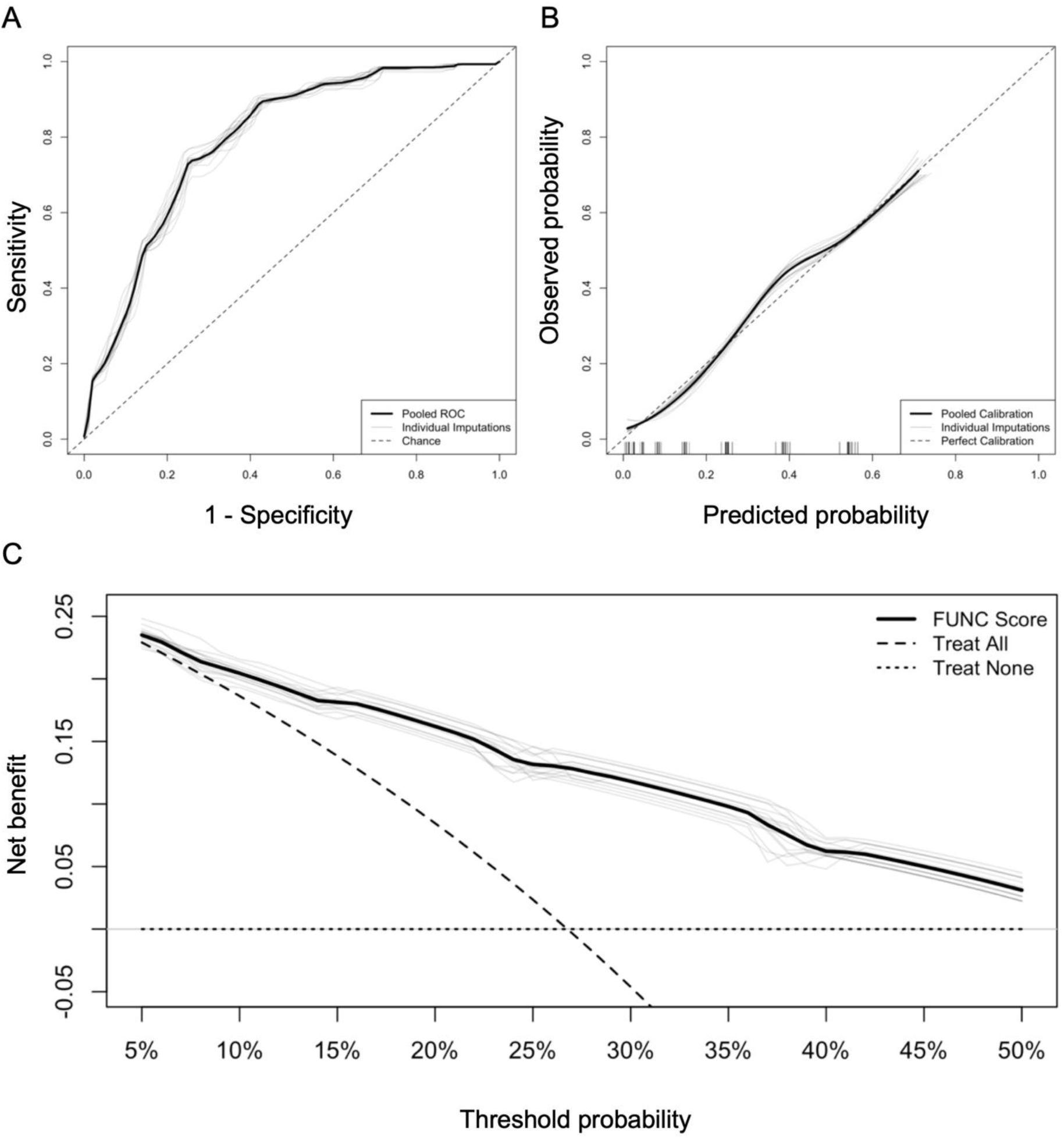
FUNC score discrimination, calibration and clinical utility. **Panel A:** Receiver operating characteristic (ROC) curve of the FUNC score for 12-month functional independence. **Panel B:** Calibration curve of the FUNC score for 12-month functional independence. **Panel C:** Decision curve analysis of the FUNC score for 12-month functional independence.

**Figure 3.**
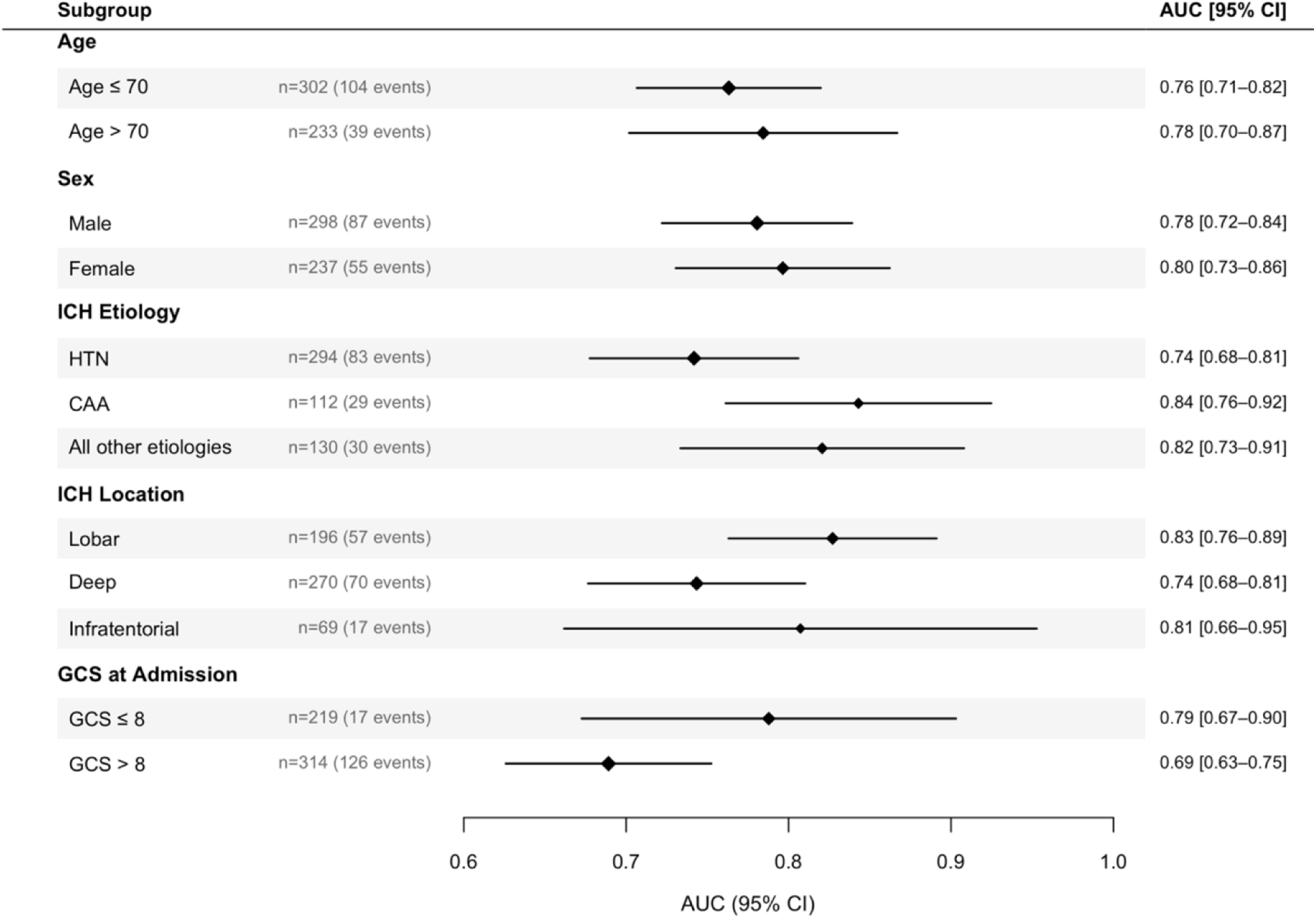
Subgroup analyses of FUNC score discrimination for 12-month functional independence. HTN denotes hypertensive microangiopathy, CAA denotes cerebral amyloid angiopathy, and GCS denotes Glasgow Coma Scale.

### Efficiency gain simulations

The distribution of FUNC-based predicted probabilities of 12-month functional independence in the complete case cohort is presented in **Figure S3**. The poor (0-6% predicted probabilities), intermediate (7-35% predicted probabilities) and good (36-100% predicted probabilities) prognosis strata were composed of 166, 186, and 93 patients, respectively. The stratum-specific thresholds that maximised equipoise were GOS ≥2, ≥3, and ≥4 for the poor, intermediate, and good prognosis strata respectively. However, since a GOS of 2 corresponds to a vegetative state, which is not considered a favorable clinical outcome irrespective of baseline prognosis, a threshold of GOS ≥3 was applied to the poor prognosis stratum, effectively collapsing it with the intermediate prognosis stratum. The sliding dichotomy thresholds for a favorable outcome were therefore GOS ≥3 for poor and intermediate prognosis strata and GOS ≥4 for the good prognosis stratum. The observed probabilities of a favorable outcome in each stratum based on the fixed versus sliding dichotomy thresholds were 3.9% versus 13.9% in the poor prognosis stratum, 22.0% versus 45.7% in the intermediate prognosis stratum, and 55.9% versus 55.9% in the good prognosis stratum. The simulated power curves for each respective treatment effect under fixed and sliding dichotomy thresholds are displayed in **Figure 4**. Under the assumptions of the simulation model and compared to the fixed dichotomy, sliding dichotomy had an efficiency gain of 27% for a treatment effect of OR=2.0, and a gain of 22% for a treatment effect of OR=2.5. The efficiency gain for a treatment effect of OR=1.5 could not be calculated in this simulation study because the fixed dichotomy did not reach 80% power despite a sample size of 1000 patients.

**Figure 4.**
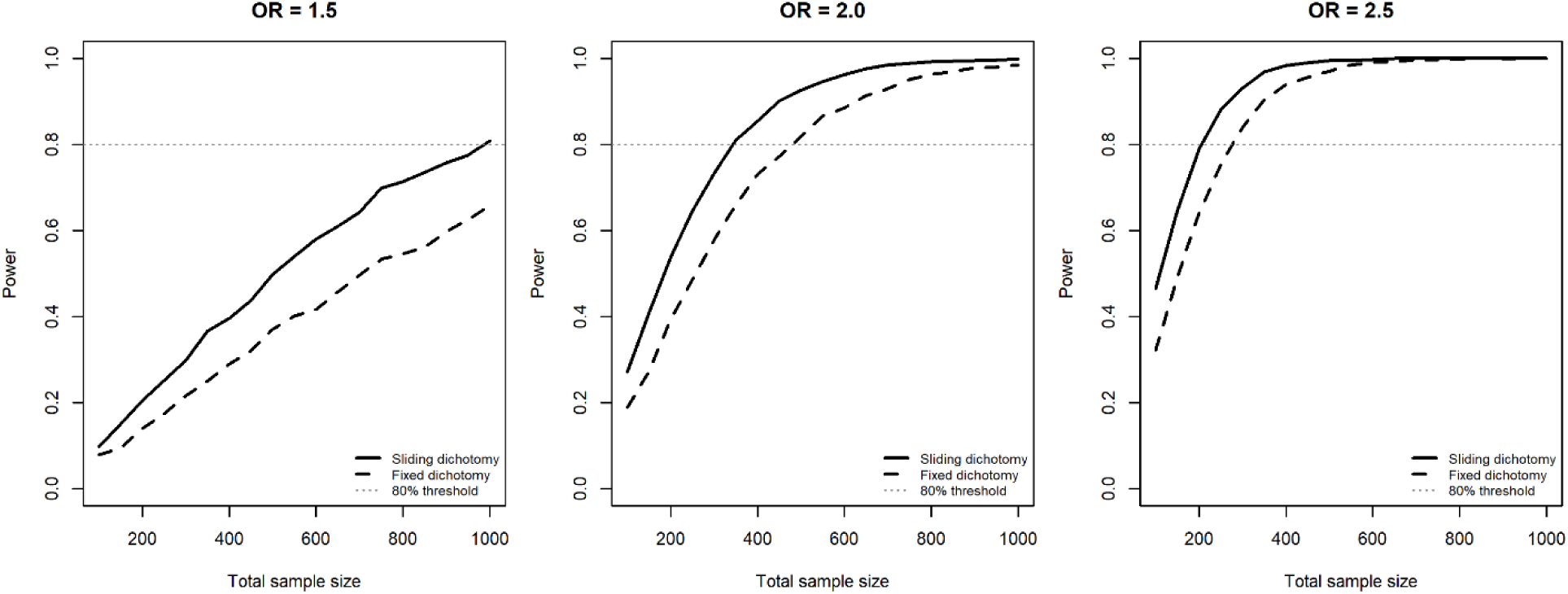
Efficiency gain simulations. OR denotes odds ratio.

## DISCUSSION

In this prospective cohort study of patients with primary ICH, the FUNC score had moderate to good discrimination and reasonable calibration for 12-month functional independence on the GOS. Its predictive accuracy was similar to the original score discrimination for 3-month functional independence (AUC of 0.82)^5^ and stable throughout the full duration of follow-up. Moreover, the score had a net benefit across clinically reasonable probability thresholds, with the greatest separation from default strategies in mid-range of threshold probabilities. These observations suggest that the FUNC score may be most informative when clinicians and family members are faced with intermediate probabilities of recovery, where distinguishing between low and moderate likelihood of functional independence is most clinically relevant.

These findings also highlight the FUNC score’s potential for improvement of outcome assessment in ICH clinical research. Most ICH prognostication scales were developed and validated to predict functional independence based on the mRS, but this scale has recognized limitations in this patient population: it overemphasizes motor function and is not tailored to the mortality and disability profiles of ICH, which are substantially different from those of acute ischemic stroke.^3, 18, 19^ There is increasing interest in using GOS and its extended form, the Glasgow Outcome Scale-Extended (GOS-E),^20, 21^ in studies of severe neurologic injuries as these scales evaluate functional status more globally and are particularly well suited for patients with impairment of consciousness.^22–24^ As such, GOS and GOS-E have been leveraged as primary^25, 26^ and secondary^9, 27^ endpoint definitions in major ICH clinical trials. Moreover, most ICH trials have relied on outcome definitions based on fixed dichotomies, which fail to consider heterogeneity in expected outcomes. Sliding dichotomy methodology, which sets different thresholds to patients based on their expected outcome, has become increasingly common in traumatic brain injury research. In this patient population, the IMPACT score predicts 12-month functional independence on GOS and GOS-E with an AUC of 0.79-0.80,^28, 29^ which enables stratified analysis where patients in different prognostic bands are evaluated against different outcome thresholds.^30, 31^ This approach increases statistical power in populations with heterogeneous outcomes by avoiding the limits of a fixed dichotomy that is easy to achieve in patients with favorable prognoses and nearly impossible in poor-prognosis patients. Exploratory simulations using FUNC-based sliding dichotomy thresholds suggested statistical efficiency gains could be significant over fixed dichotomies: the sample size reduction estimates of 20-30% were in line with gains observed in traumatic brain injury research. Provided additional external validation confirms its prognostic performance across centers, future ICH clinical trials could therefore leverage the FUNC score to take advantage of sliding dichotomy methodology.

Our study’s strengths include prospective and consecutive enrollment of a large, representative sample of patients with primary ICH, a high 12-month retention rate, use of robust imputation methodology for missing data and investigation of accuracy in clinically relevant subgroups. The 17% missing 12-month functional outcome proportion in this study is in line with observations from prior ICH cohort studies, including the original study that developed and validated the FUNC score for 3-month functional independence.^5, 32–34^ We leveraged multiple imputation by chained equations in this study because patients lost to follow up had more favorable baseline characteristics, suggesting that complete case analysis may be biased. This is an improvement over the original FUNC study that had assumed data missingness was completely at random. Our study also has limitations. First, treating clinicians were not blinded to FUNC score variable data. If these variables influenced management, including WLST, this could have led to overestimation of the score’s discrimination accuracy. However, it is not clinical practice at our institution to routinely calculate or use the FUNC score to guide management, which is congruent with recent guidelines on prognostication after ICH.^2^ Furthermore, the observed overestimation of functional independence at low predicted probabilities suggests the FUNC score may provide overly optimistic prognosis for the most severely affected patients – a miscalibration pattern that would act against premature WLST rather than facilitate it, partially mitigating concerns about a self-fulfilling prophecy of poor outcome. Second, this study computed cross-validated estimates within the same cohort, so these estimates represent internal validity only and warrant additional external validation prior to widespread use for family counseling and clinical research. Third, subgroup analyses were underpowered to detect heterogeneity in FUNC score prognostic accuracy. Future work should investigate whether this score is similarly applicable to both lobar and deep ICH, conditions in which the prognostic implications of age and hematoma volume may be different.^35–37^ Fourth, simulated efficiency gains were calculated based on several simplifying assumptions: perfect prognostic classification at trial entry, certainty in favorable outcome probabilities, absence of bias in favorable outcome probability estimates, and homogeneity in treatment effect within each stratum. In real world contexts, FUNC score misclassification and variability in FUNC score prognostic accuracy could attenuate the observed efficiency gain. Moreover, the efficiency gain in trials of interventions that aim to decrease the incidence of extreme outcomes such as death might be smaller than those obtained through this simulation. Finally, because the poor and intermediate strata shared a common threshold, the framework evaluated here approximates a two-tier rather than a fully three-tier sliding dichotomy. Fifth, findings are not generalizable to institutions with different resources or medical practices, as well as the pediatric patient population.

In conclusion, the FUNC score has moderate to good predictive accuracy for functional independence 12 months after primary ICH. Provided additional external validation confirms its prognostic performance across centers, this GOS-based score may be leveraged in ICH clinical research to take advantage of sliding dichotomy methodology and be helpful to counsel families on global measures of functional independence in the long term.

## Data Availability

The data and code that support the findings of this study are available from the corresponding author upon reasonable request.

## ACKNOWLEDGMENTS

The authors express gratitude to all ICHOP investigators and collaborators.

## CONTRIBUTIONS

JNB, QS and JC conceptualized the study. VK, YLS, and ABC collected data. JNB performed data analysis and drafted the manuscript. All other authors revised the manuscript for intellectual content. All authors reviewed and approved the final version of the manuscript. JC is the senior author and the guarantor of the study.

## ABBREVIATIONS AND ACRONYMS

AUC: Area under the receiver operating characteristic curve
FUNC: Functional Outcome in Patients with Primary Intracerebral Hemorrhage
GCS: Glasgow Coma Scale
GOS: Glasgow Outcome Scale
GOS-E: Glasgow Outcome Scale - Extended
ICH: Intracerebral hemorrhage
ICHOP: Intracerebral Hemorrhage Outcomes Project
mRS: Modified Rankin Scale
NIHSS: National Institutes of Health Stroke Scale
OR: Odds ratio
WLST: Withdrawal of life-sustaining therapies

## REFERENCES

1. Greenberg SM, Ziai WC, Cordonnier C, Dowlatshahi D, Francis B, Goldstein JN, et al. 2022 Guideline for the Management of Patients With Spontaneous Intracerebral Hemorrhage: A Guideline From the American Heart Association/American Stroke Association. Stroke. 2022;53(7):e282–e361.

2. Hwang DY, Kim KS, Muehlschlegel S, Wartenberg KE, Rajajee V, Alexander SA, et al. Guidelines for Neuroprognostication in Critically Ill Adults with Intracerebral Hemorrhage. Neurocrit Care. 2024;40(2):395–414.

3. Witsch J, Siegerink B, Nolte CH, Sprugel M, Steiner T, Endres M, et al. Prognostication after intracerebral hemorrhage: a review. Neurol Res Pract. 2021;3(1):22.

4. Gregorio T, Pipa S, Cavaleiro P, Atanasio G, Albuquerque I, Chaves PC, et al. Assessment and Comparison of the Four Most Extensively Validated Prognostic Scales for Intracerebral Hemorrhage: Systematic Review with Meta-analysis. Neurocrit Care. 2019;30(2):449–66.

5. Rost NS, Smith EE, Chang Y, Snider RW, Chanderraj R, Schwab K, et al. Prediction of Functional Outcome in Patients With Primary Intracerebral Hemorrhage. Stroke. 2008;39(8):2304–9.

6. Mai LM, Joundi RA, Katsanos AH, Selim M, Shoamanesh A. Pathophysiology of Intracerebral Hemorrhage: Recovery Trajectories. Stroke. 2025;56(3):783–93.

7. Morotti A, Nawabi J, Pilotto A, Toffali M, Busto G, Mazzacane F, et al. Functional outcome improvement from 3 to 12 months after intracerebral hemorrhage. Eur Stroke J. 2024;9(2):391–7.

8. Arthur AS, Jahromi BS, Saphier PS, Nickele CM, Ryan RW, Vajkoczy P, et al. Minimally Invasive Surgery vs Medical Management Alone for Intracerebral Hemorrhage: The MIND Randomized Clinical Trial. JAMA Neurol. 2025;82(11):1113–21.

9. Hanley DF, Thompson RE, Rosenblum M, Yenokyan G, Lane K, McBee N, et al. Efficacy and safety of minimally invasive surgery with thrombolysis in intracerebral haemorrhage evacuation (MISTIE III): a randomised, controlled, open-label, blinded endpoint phase 3 trial. Lancet. 2019;393(10175):1021–32.

10. Pradilla G, Ratcliff JJ, Hall AJ, Saville BR, Allen JW, Paulon G, et al. Trial of Early Minimally Invasive Removal of Intracerebral Hemorrhage. N Engl J Med. 2024;390(14):1277–89.

11. Gupta VP, Garton ALA, Sisti JA, Christophe BR, Lord AS, Lewis AK, et al. Prognosticating Functional Outcome After Intracerebral Hemorrhage: The ICHOP Score. World Neurosurg. 2017;101:577–83.

12. Collins GS, Moons KGM, Dhiman P, Riley RD, Beam AL, Van Calster B, et al. TRIPOD+AI statement: updated guidance for reporting clinical prediction models that use regression or machine learning methods. BMJ. 2024;385:e078378.

13. Bodner TE. What Improves with Increased Missing Data Imputations? Structural Equation Modeling: A Multidisciplinary Journal. 2008;15(4):651–75.

14. White IR, Royston P, Wood AM. Multiple imputation using chained equations: Issues and guidance for practice. Stat Med. 2011;30(4):377–99.

15. Rubin DB. Multiple Imputation After 18+ Years. Journal of the American Statistical Association. 1996;91(434):473–89.

16. Rufibach K. Use of Brier score to assess binary predictions. J Clin Epidemiol. 2010;63(8):938–9.

17. Vickers AJ, Elkin EB. Decision curve analysis: a novel method for evaluating prediction models. Med Decis Making. 2006;26(6):565–74.

18. Banks JL, Marotta CA. Outcomes Validity and Reliability of the Modified Rankin Scale: Implications for Stroke Clinical Trials. Stroke. 2007;38(3):1091–6.

19. Braun RG, Heitsch L, Cole JW, Lindgren AG, de Havenon A, Dude JA, et al. Domain-Specific Outcomes for Stroke Clinical Trials: What the Modified Rankin Isn’t Ranking. Neurology. 2021;97(8):367–77.

20. Jennett B, Snoek J, Bond MR, Brooks N. Disability after severe head injury: observations on the use of the Glasgow Outcome Scale. J Neurol Neurosurg Psychiatry. 1981;44(4):285–93.

21. Wilson JT, Pettigrew LE, Teasdale GM. Structured interviews for the Glasgow Outcome Scale and the extended Glasgow Outcome Scale: guidelines for their use. J Neurotrauma. 1998;15(8):573–85.

22. Chen KY, Kung WM, Kuo LT, Huang AP. Ultrarapid Endoscopic-Aided Hematoma Evacuation in Patients with Thalamic Hemorrhage. Behav Neurol. 2021;2021:8886004.

23. Fatima N, Razzaq S, El Beltagi A, Shuaib A, Saqqur M. Decompressive Craniectomy: A Preliminary Study of Comparative Radiographic Characteristics Predicting Outcome in Malignant Ischemic Stroke. World Neurosurg. 2020;133:e267–e74.

24. Stienen MN, Visser-Meily JM, Schweizer TA, Hänggi D, Macdonald RL, Vergouwen MDI. Prioritization and Timing of Outcomes and Endpoints After Aneurysmal Subarachnoid Hemorrhage in Clinical Trials and Observational Studies: Proposal of a Multidisciplinary Research Group. Neurocrit Care. 2019;30(Suppl 1):102–13.

25. Mendelow AD, Gregson BA, Fernandes HM, Murray GD, Teasdale GM, Hope DT, et al. Early surgery versus initial conservative treatment in patients with spontaneous supratentorial intracerebral haematomas in the International Surgical Trial in Intracerebral Haemorrhage (STICH): a randomised trial. Lancet. 2005;365(9457):387–97.

26. Mendelow AD, Gregson BA, Rowan EN, Murray GD, Gholkar A, Mitchell PM. Early surgery versus initial conservative treatment in patients with spontaneous supratentorial lobar intracerebral haematomas (STICH II): a randomised trial. Lancet. 2013;382(9890):397–408.

27. Hanley DF, Lane K, McBee N, Ziai W, Tuhrim S, Lees KR, et al. Thrombolytic removal of intraventricular haemorrhage in treatment of severe stroke: results of the randomised, multicentre, multiregion, placebo-controlled CLEAR III trial. Lancet. 2017;389(10069):603–11.

28. Eagle SR, Nwachuku E, Elmer J, Deng H, Okonkwo DO, Pease M. Performance of CRASH and IMPACT Prognostic Models for Traumatic Brain Injury at 12 and 24 Months Post-Injury. Neurotrauma Rep. 2023;4(1):118–23.

29. Rostami E, Gustafsson D, Hånell A, Howells T, Lenell S, Lewén A, et al. Prognosis in moderate-severe traumatic brain injury in a Swedish cohort and external validation of the IMPACT models. Acta Neurochir (Wien). 2022;164(3):615–24.

30. Steyerberg EW, Mushkudiani N, Perel P, Butcher I, Lu J, McHugh GS, et al. Predicting outcome after traumatic brain injury: development and international validation of prognostic scores based on admission characteristics. PLoS Med. 2008;5(8):e165; discussion e.

31. Yeatts SD, Martin RH, Meurer W, Silbergleit R, Rockswold GL, Barsan WG, et al. Sliding Scoring of the Glasgow Outcome Scale-Extended as Primary Outcome in Traumatic Brain Injury Trials. J Neurotrauma. 2020;37(24):2674–9.

32. Abulhasan YB, Teitelbaum J, Al-Ramadhani K, Morrison KT, Angle MR. Functional Outcomes and Mortality in Patients With Intracerebral Hemorrhage After Intensive Medical and Surgical Support. Neurology. 2023;100(19):e1985–e95.

33. Ziai W, Woo D, Sansing L, Hanley D, Ostapkovich N, Triene K, et al. The REpeated ASSEssment of SurvivorS in intracerebral haemorrhage: protocol for a multicentre, prospective observational study. BMJ Open. 2025;15(2):e094322.

34. Appelboom G, Bruce SS, Han J, Piazza M, Hwang B, Hickman ZL, et al. Functional outcome prediction following intracerebral hemorrhage. Journal of Clinical Neuroscience. 2012;19(6):795–8.

35. Delcourt C, Sato S, Zhang S, Sandset EC, Zheng D, Chen X, et al. Intracerebral hemorrhage location and outcome among INTERACT2 participants. Neurology. 2017;88(15):1408–14.

36. Kuohn LR, Witsch J, Steiner T, Sheth KN, Kamel H, Navi BB, et al. Early Deterioration, Hematoma Expansion, and Outcomes in Deep Versus Lobar Intracerebral Hemorrhage: The FAST Trial. Stroke. 2022;53(8):2441–8.

37. Roh D, Boehme A, Young C, Roth W, Gutierrez J, Flaherty M, et al. Hematoma expansion is more frequent in deep than lobar intracerebral hemorrhage. Neurology. 2020;95(24):e3386–e93.

