## Supplemental Materials for "Extension of the FUNC score for prediction of 12-month functional independence after primary intracerebral hemorrhage"

### SUPPLEMENTAL MATERIAL

#### TABLE OF CONTENT


Supplemental Figures

Figure S1 p. 2

Figure S2 p. 3

Figure S3 p. 4

Supplemental Tables

Table S1 p. 5

Table S2 p. 6

### SUPPLEMENTAL FIGURES

**Figure S1.** Probabilities of 12-month functional independence across FUNC score bins.

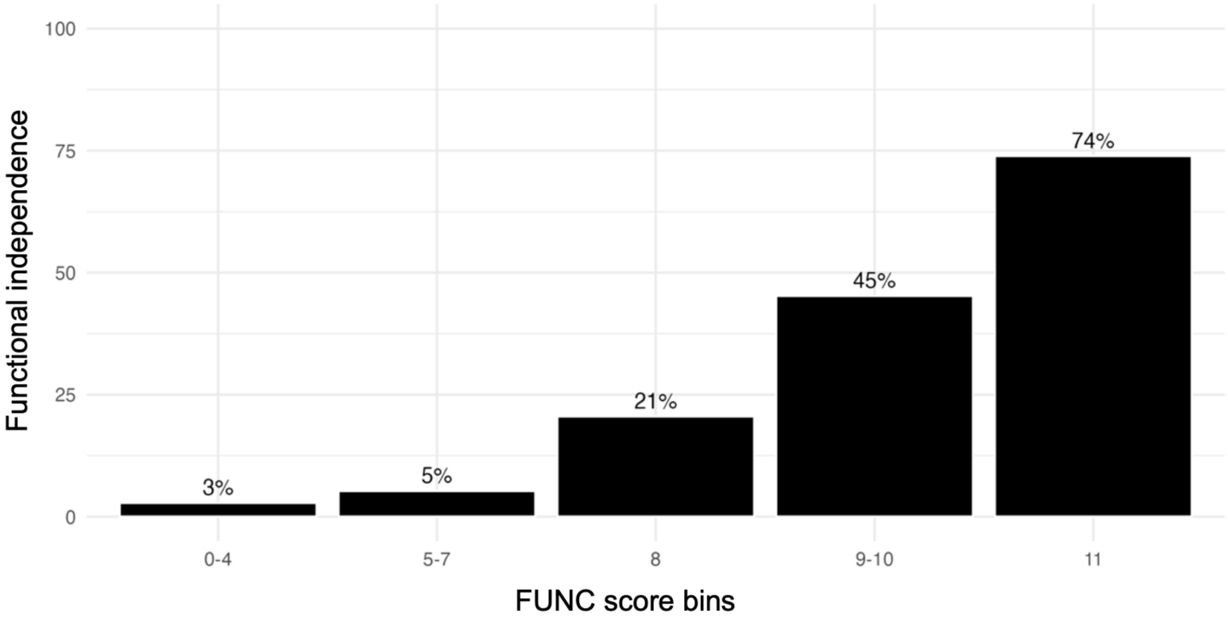

**Figure S2.** FUNC score discrimination and calibration in sensitivity complete case analysis.

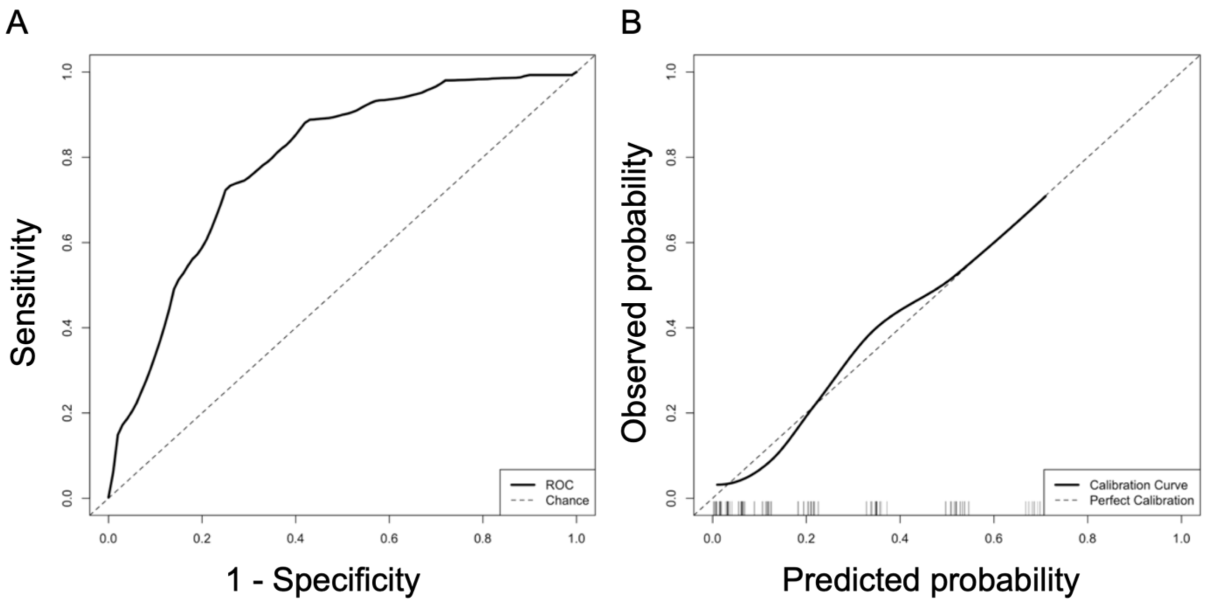


**Panel A:** Receiver operating characteristics (ROC) curve of the FUNC score for 12-month functional independence. **Panel B:** Calibration curve of the FUNC score for 12-month functional independence.

**Figure S3.** Distribution of FUNC-based predicted probabilities for 12-month functional independence.

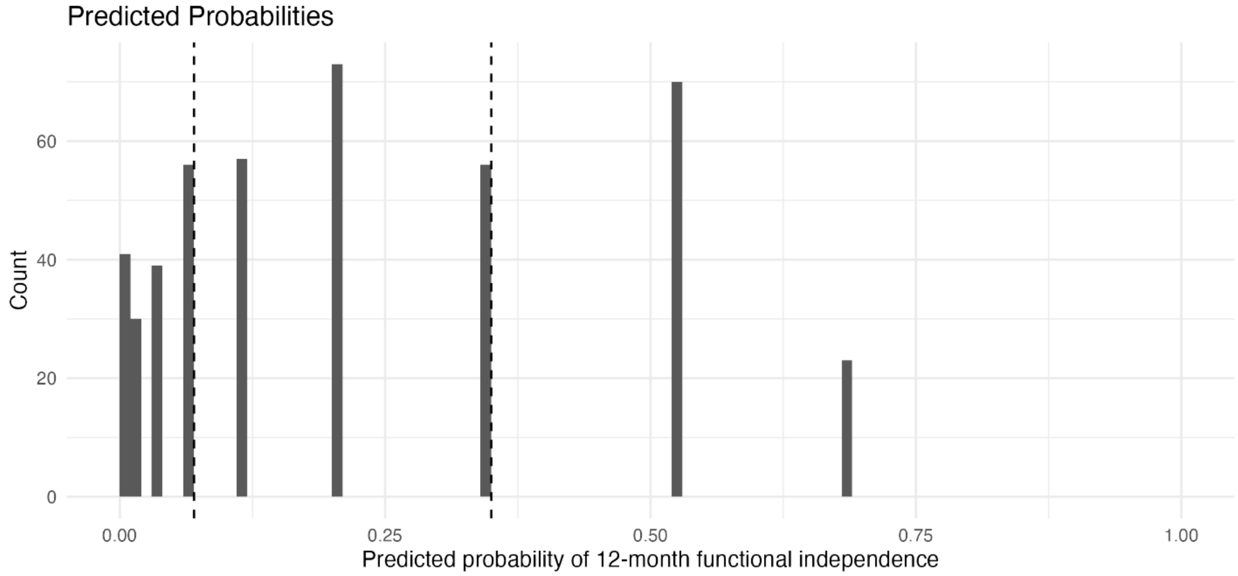


The vertical dashed lines represent the boundaries of the three prognostic strata. The probabilities of 12-month functional independence ranged from 0-6% for the poor prognosis stratum, 7-35% for the intermediate prognosis stratum, and 36-100% for the good prognosis stratum.

### SUPPLEMENTAL TABLES

**Table S1**. FUNC score components.

| Variable | Points |
|---|---|
| Age | |
| < 70 | 2 |
| 70-79 | 1 |
| ≥ 80 | 0 |
| Pre-ICH cognitive impairment | |
| No | 1 |
| Yes | 0 |
| ICH volume, $cm^3$ | |
| < 30 | 4 |
| 30-60 | 2 |
| > 60 | 0 |
| ICH location | |
| Lobar | 2 |
| Deep | 1 |
| Infratentorial | 0 |
| GCS score | |
| ≥ 9 | 2 |
| ≤ 8 | 0 |

FUNC denotes Functional Outcome in Patients with Primary Intracerebral Hemorrhage, ICH denotes intracerebral hemorrhage and GCS denotes Glasgow Coma Scale. Reference: Rost NS, Smith EE, Chang Y, Snider RW, Chanderraj R, Schwab K, et al. Prediction of Functional Outcome in Patients With Primary Intracerebral Hemorrhage. *Stroke*. 2008;39(8):2304-9.

**Table S2**. Characteristics and in-hospital outcomes of patients with and without 12-mo GOS.

| | Patients with 12-month GOS (n=445) | Patients without 12-month GOS (n=90) | Absolute SMD |
|---|---|---|---|
| **Demographics** | | | |
| Median age (IQR), years | 69 (55-79) | 61 (52-76) | 0.20 |
| Female sex, n (%) | 196 (44) | 41 (46) | 0.03 |
| **Clinical data at hospital admission** | | | |
| Median premorbid mRS (IQR) | 0 (0-2) | 0 (0-1) | 0.32 |
| History of dementia, n (%) | 43 (10) | 4 (4) | 0.21 |
| Median NIHSS (IQR) | 17 (6-26) | 10 (4-17) | 0.54 |
| Median GCS (IQR) | 9 (5-14) | 14 (10-15) | 0.64 |
| ICH etiology, n (%) [a] | | | 0.74 |
| Hypertensive microangiopathy | 232 (52) | 62 (69) | |
| Cerebral amyloid angiopathy | 95 (21) | 17 (19) | |
| Anticoagulation | 61 (14) | 2 (2) | |
| Coagulopathy | 24 (5) | 2 (2) | |
| Drug induced | 1 (0.2) | 2 (2) | |
| Other | 22 (5) | 3 (3) | |
| Unknown | 18 (4) | 7 (8) | |
| **Neuroimaging data at first CT scan** | | | |
| Localization of ICH, n (%) | | | 0.12 |
| Lobar | 164 (37) | 32 (36) | |
| Deep | 221 (50) | 49 (54) | |
| Infratentorial | 60 (13) | 9 (10) | |
| Median hematoma volume (IQR), mL | 18 (6-44) | 12 (5-28) | 0.03 |
| Intraventricular hemorrhage, n (%) | 223 (50) | 47 (52) | 0.04 |
| **Admission FUNC** | | | |
| Median FUNC (IQR) | 7 (6-9) | 9 (8-10) | 0.74 |
| **Outcomes during hospitalization** | | | |
| Median hospital length of stay (IQR), days | 8 (4-18) | 10 (6-21) | 0.16 |
| Discharge disposition, n (%) | | | 1.20 |
| Home, with or without outpatient rehabilitation | 63 (14) | 15 (17) | |
| Inpatient rehabilitation facility | 122 (28) | 52 (58) | |
| Skilled nursing facility | 86 (19) | 18 (20) | |
| Other hospital | 13 (3) | 4 (4) | |
| Other location | | | |
| Death | 167 (38) | 1 (1) | |
| Unknown | 2 (0.4) | 0 (0) | |
| WLST, n (%) | 64 (14) | 0 (0) | 0.58 |
| Median discharge mRS (IQR) | 5 (4-6) | 4 (3-4) | 0.77 |

Notes: [a] Not mutually exclusive categories. SMD denotes standardized mean difference, IQR denotes interquartile range, mRS denotes modified Rankin Scale, NIHSS denotes National Institutes of Health Stroke Scale, GCS denotes Glasgow Coma Scale, ICH denotes intracerebral hemorrhage, FUNC denotes Functional Outcome in Patients with Primary Intracerebral Hemorrhage, and WLST denotes withdrawal of life-sustaining therapies.
